# The association between high ambient temperature and mortality in the Mediterranean basin: a systematic review and meta-analysis

**DOI:** 10.1101/2022.01.20.22269580

**Authors:** Talila Perry, Uri Obolski, Chava Peretz

## Abstract

**Background:** The effect of hot ambient temperature on mortality differs between geographical locations. The Mediterranean basin has been identified as one of the most prominent “hot-spots” in the world in terms of vulnerability to climate change. No recent, large synthesis of the estimated effects in the Mediterranean basin has been conducted.

**Methods:** A systematic review was conducted across three major databases to retrieve effect estimates from time-series or case-crossover studies on temperature and mortality performed in the Mediterranean basin countries, between 2000-2021. Of all reviewed studies, n=16 were eligible for inclusion in the meta-analysis. Estimates were pooled using a random-effects model. Subgroup analyses were performed for death-cause, age-group, region, and climate type. Meta-regression was performed with respect to national Gross Domestic Product (GDP) per capita, latitude and longitude, and local temperature thresholds.

**Results:** We found an increased risk of all-cause mortality due to ambient heat exposure in the Mediterranean basin, with a pooled RR=1.035 (95%CI 1.028-1.041) per 1°C increase in temperature above local thresholds (I^2^=79%). Risk was highest for respiratory (RR=1.063, 95% CI 1.052-1.074) and cardiovascular (RR=1.046, 95% CI 1.036-1.057) mortality.

**Conclusions:** Hot ambient temperatures increase the mortality risk across the Mediterranean basin. This is increasingly important for public health processes in the Mediterranean basin countries in light of the climate changes already noticed in this area. Further high-quality studies, especially in North African, Asian Mediterranean, and eastern European countries, are needed to bolster regional preparedness against future heat-related health burdens.

**Key Messages:**

- High ambient temperatures affect short-term mortality across the Mediterranean basin countries.
- Risk is highest for respiratory and cardiovascular mortality.
- In light of climate change, this is an increasingly important public health concern. Further high-quality studies, especially in North African, Asian Mediterranean and eastern European countries, are needed to help regions prepare for future heat-related health burdens.

## Introduction

Climate change, and especially global warming, is a matter of increasing public health concern due to the adverse associations between heat and increased mortality(1,2). Time-series studies on the short-term relationships between ambient temperature and daily mortality reported the dose-response curve to be generally “J”-shaped, with a sharper increase in mortality risk above a regional-specific threshold temperature(3–5). Heat-induced mortality across and within regions depends strongly on local climate and geographical features. The Mediterranean basin has been identified as one of the world’s most prominent “hot-spots” in terms of vulnerability to climate change(6). A pronounced decrease in precipitation, as well as increasing mean temperatures and warm-season variability, make the region highly responsive to even slight meteorological changes(7). Extreme and uncharacteristic hot weather events have already been observed in the Mediterranean basin, particularly in southwest Europe, the Iberian peninsula, and southern France(1).

Socio-demography also plays a significant role in population vulnerability to heat. Specifically, urbanisation, older age, population density, poor health status, lower socioeconomic status, and cultural patterns, such as decreased social contacts, have been shown to increase heat-related mortality(8). Accordingly, Europe exhibits a high level of urbanisation, and projections put 34% of its population above 60 years old by 2050(9). This trend of aging and urbanisation of European populations may further increase the risk of heat-induced mortality in this region.

The extent of the risk, however, is still unclear. Epidemiological studies analysing Mediterranean countries have predominantly focused on high-income European countries. Hence, regions expected to experience the most severe consequences from heat waves, such as Asian and African countries in the Mediterranean basin, are underrepresented in research(10).

To the best of our knowledge, no recent, large synthesis of the estimated heat related mortality risks of such regions has been conducted. Therefore, we present here a systematic review and meta-analysis of the literature on hot temperatures and mortality in the Mediterranean basin countries, and pool estimates from European and non-European Mediterranean countries to calculate the effect of short-term exposure to high ambient temperatures on mortality. In addition, we investigate the heterogeneity resulting from methodological, geographic, and sociodemographic differences in the collected studies.

## Methods

This systematic review and meta-analysis was performed in accordance with the Preferred Reporting Items for Systematic Reviews and Meta-Analyses (PRISMA) statement and criteria(11).

### Inclusion criteria

The studies selected for inclusion were epidemiological time-series or case-crossover studies on ambient heat (measured as daily temperature or an index combining temperature and humidity) and mortality outcomes, in countries with a Mediterranean coast. The peer reviewed databases of Ovid Medline, Embase, and Web of Science were searched for papers published between 1 January 2000 and 17 February 2021. Terms included in the search were: heat, temperature, weather, climate, and mortality, as well the names of the different Mediterranean countries.References cited in the included studies and relevant review articles were also searched.

1. Outcome was defined as daily mortality, measured as all-cause mortality, natural non-external mortality, or mortality from specific causes (e.g. respiratory or cardiovascular causes).
2. Quantitative estimates of ambient heat-related mortality given as added risk per 1°C above a location-specific threshold were used.

If studies reported the use of the same dataset, only the most recent study was included.

### Data collection

Qualitative and quantitative characteristics of the studies with relative risks (RR) and 95% confidence intervals (CI), were recorded. The RR of the largest spatial unit reported in each study was considered, as well as the RRs for vulnerable subgroups.

### Statistical analysis

#### Meta-analysis

A random effects model was used to estimate the RRs of mortality per 1°C increase in temperature above a local threshold for the entire Mediterranean basin. For multi-center studies reporting estimates for multiple sites in the same or several countries, the site effects were first pooled by applying a random effects model, and then the pooled estimates were input into the overall meta-analysis. Heterogeneity between studies was determined using the I^2^ statistic. Influence analyses were conducted with the help of a Baujat plot to test for outlying studies and possible reduction of heterogeneity.

#### Subgroup meta-analyses

Subgroup meta-analyses were performed to reduce heterogeneity.

This involved, subgrouping and analyses by cardiovascular or respiratory death-causes, followed by subgrouping and analyses by age. Populations > 65 years (if provided) were analysed separately. In studies with RRs of discrete age groups above 65, a random effects meta-analysis was used to pool the estimates into a site-specific over 65 estimate. For studies with multiple sites, the appropriate site-specific estimates were pooled to yield the overall over 65 estimate.

Subgroup analyses were performed on the smallest spatial unit available from the data, regardless of the study in which they appeared (Supplementary Table S2). These spatial units were pooled by:

1. Geographical location – all spatial units were stratified to three regions: western Europe, eastern Europe, and the Middle East and North Africa (MENA). Istanbul was considered a part of eastern Europe, and a sensitivity analysis was performed with Istanbul in the MENA region.
2. City-specific climate -all spatial units were stratified by their climate type based on the widely used Köppen-Geiger climate classification(12) (Supplementary Table S1).

Additional subgroup analyses by type of statistical model chosen, seasonality (all-year or warm months), and length of time series (less than 10 years, or 10 years and more) were carried out as sensitivity analyses.

### Meta-regression

Meta regression analyses were performed to further explore the factors affecting heat related mortality and address between-study heterogeneity using the following continuous variables:

1. Gross domestic product (GDP) per capita of the country containing the city/region, using the most recent estimate from the World Bank(13).
2. Latitude and longitude of cities (or center of region or country if the unit is larger than a city).
3. Study publication year, to detect whether the effects of heat have changed over time.
4. Site-specific heat thresholds.

All statistical analyses were carried out using the *meta* package of R version 4.1.1 in Rstudio version 2021.09.0(14).

## Results

The systematic search yielded 4941 articles. After removal of duplicates and irrelevant articles, 124 full-text papers were read, and 16 papers met the inclusion criteria (Figure 1).

**Figure 1:**
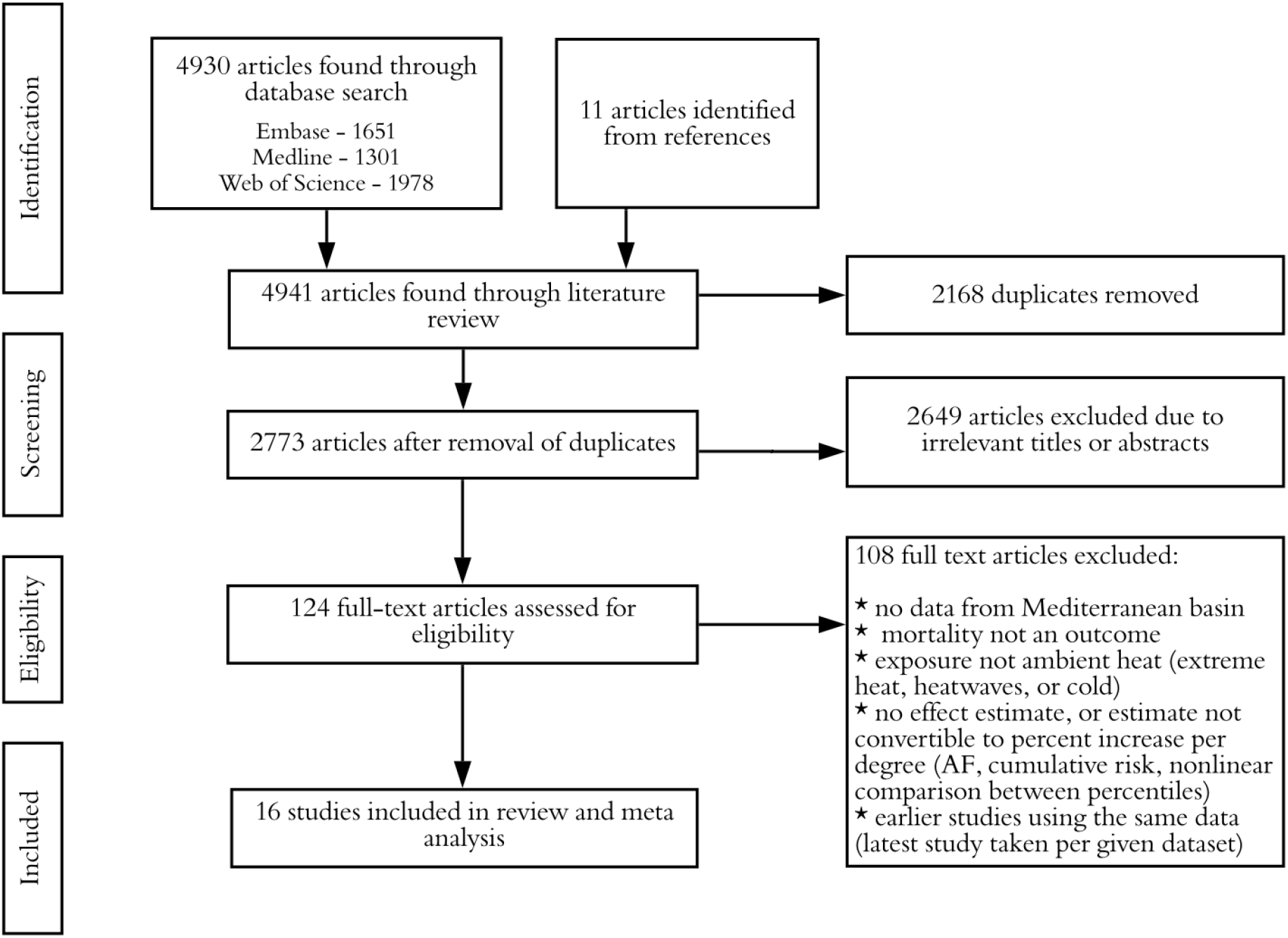
Flow chart of inclusion and exclusion of papers following PRISMA guidelines

### Characteristics of included studies

All 16 included studies were published between 1988 and 2012, with durations ranging from 3 to 21 years, although most (*n*=12) were shorter than 10 years. Ten Mediterranean countries were studied, six in Europe (Cyprus, Greece, Italy, Portugal, Slovenia, and Spain), three in Asia (Israel, Lebanon, and Turkey), and one in North Africa (Tunisia). Most of the studied locations have a temperate climate (Supplementary Tables S1 and S2). Measurements of ambient heat included the mean (*n*=6) or maximum (*n*=4) daily temperatures, or indices combining temperature and humidity: maximum apparent temperature (*n*=4); discomfort index (DI, one study); and universal thermal climate index (UTCI, one study; Table 1). The outcome of daily mortality was defined as natural or non-external (*n*=11), all-cause mortality (*n*=4), or death due to cardiovascular or respiratory causes only (one study; Table 1). Nine studies provided risk estimates for both overall mortality and for the over 65 years old population. Six studies provided specific estimates for cardiovascular causes of death, and five studies for respiratory causes (Table 1).

**Table 1:**
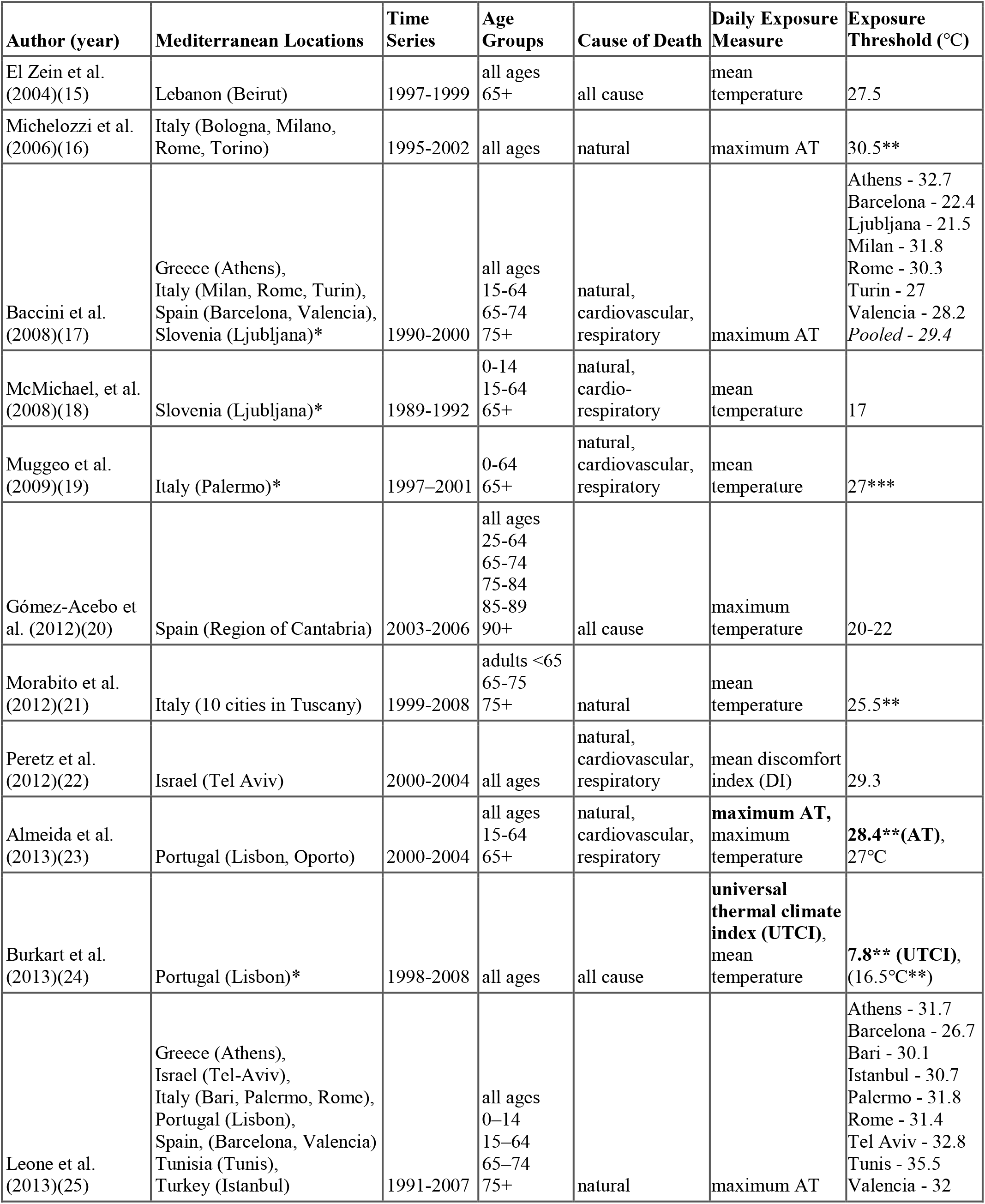
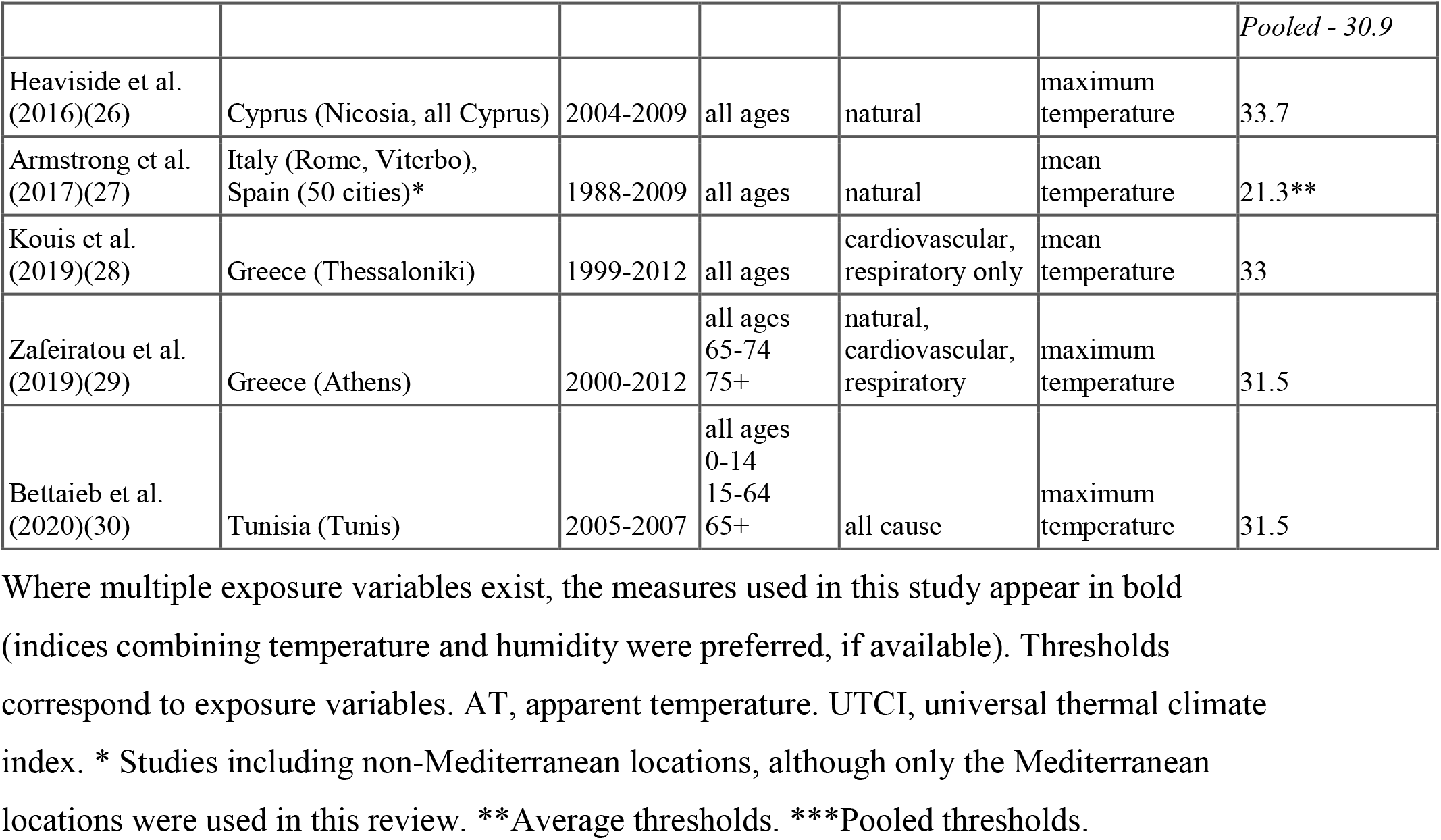
Study characteristics

| Author (year) | Mediterranean Locations | Time Series | Age Groups | Cause of Death | Daily Exposure Measure | Exposure Threshold (°C) |
| --- | --- | --- | --- | --- | --- | --- |
| El Zein et al. (2004)(15) | Lebanon (Beirut) | 1997-1999 | all ages<br>65+ | all cause | mean temperature | 27.5 |
| Michelozzi et al. (2006)(16) | Italy (Bologna, Milano, Rome, Torino) | 1995-2002 | all ages | natural | maximum AT | 30.5** |
| Baccini et al. (2008)(17) | Greece (Athens), Italy (Milan, Rome, Turin), Spain (Barcelona, Valencia), Slovenia (Ljubljana)* | 1990-2000 | all ages<br>15-64<br>65-74<br>75+ | natural, cardiovascular, respiratory | maximum AT | Athens - 32.7<br>Barcelona - 22.4<br>Ljubljana - 21.5<br>Milan - 31.8<br>Rome - 30.3<br>Turin - 27<br>Valencia - 28.2<br>*Pooled* - 29.4 |
| McMichael, et al. (2008)(18) | Slovenia (Ljubljana)* | 1989-1992 | 0-14<br>15-64<br>65+ | natural, cardio-respiratory | mean temperature | 17 |
| Muggeo et al. (2009)(19) | Italy (Palermo)* | 1997–2001 | 0-64<br>65+ | natural, cardiovascular, respiratory | mean temperature | 27*** |
| Gómez-Acebo et al. (2012)(20) | Spain (Region of Cantabria) | 2003-2006 | all ages<br>25-64<br>65-74<br>75-84<br>85-89<br>90+ | all cause | maximum temperature | 20-22 |
| Morabito et al. (2012)(21) | Italy (10 cities in Tuscany) | 1999-2008 | adults <65<br>65-75<br>75+ | natural | mean temperature | 25.5** |
| Peretz et al. (2012)(22) | Israel (Tel Aviv) | 2000-2004 | all ages | natural, cardiovascular, respiratory | mean discomfort index (DI) | 29.3 |
| Almeida et al. (2013)(23) | Portugal (Lisbon, Oporto) | 2000-2004 | all ages<br>15-64<br>65+ | natural, cardiovascular, respiratory | **maximum AT**, maximum temperature | **28.4\*\* (AT)**, 27°C |
| Burkart et al. (2013)(24) | Portugal (Lisbon)* | 1998-2008 | all ages | all cause | **universal thermal climate index (UTCI)**, mean temperature | **7.8\*\* (UTCI)**, (16.5°C**) |
| Leone et al. (2013)(25) | Greece (Athens), Israel (Tel-Aviv), Italy (Bari, Palermo, Rome), Portugal (Lisbon), Spain, (Barcelona, Valencia) Tunisia (Tunis), Turkey (Istanbul) | 1991-2007 | all ages<br>0–14<br>15–64<br>65–74<br>75+ | natural | maximum AT | Athens - 31.7<br>Barcelona - 26.7<br>Bari - 30.1<br>Istanbul - 30.7<br>Palermo - 31.8<br>Rome - 31.4<br>Tel Aviv - 32.8<br>Tunis - 35.5<br>Valencia - 32 |
|  |  |  |  |  |  | *Pooled - 30.9* |
| Heaviside et al.<br>(2016)(26) | Cyprus (Nicosia, all Cyprus) | 2004-2009 | all ages | natural | maximum temperature | 33.7 |
| Armstrong et al.<br>(2017)(27) | Italy (Rome, Viterbo),<br>Spain (50 cities)* | 1988-2009 | all ages | natural | mean temperature | 21.3** |
| Kouis et al.<br>(2019)(28) | Greece (Thessaloniki) | 1999-2012 | all ages | cardiovascular,<br>respiratory only | mean temperature | 33 |
| Zafeiratou et al.<br>(2019)(29) | Greece (Athens) | 2000-2012 | all ages<br>65-74<br>75+ | natural,<br>cardiovascular,<br>respiratory | maximum temperature | 31.5 |
| Bettaieb et al.<br>(2020)(30) | Tunisia (Tunis) | 2005-2007 | all ages<br>0-14<br>15-64<br>65+ | all cause | maximum temperature | 31.5 |

Where multiple exposure variables exist, the measures used in this study appear in bold (indices combining temperature and humidity were preferred, if available). Thresholds correspond to exposure variables. AT, apparent temperature. UTCI, universal thermal climate index. \* Studies including non-Mediterranean locations, although only the Mediterranean locations were used in this review. \*\*Average thresholds. \*\*\*Pooled thresholds.

Other methodological and modelling differences between the studies included seasonality, statistical models used, exposure lags, and confounder adjustment (Supplementary Table S3).

### The heat-mortality relationship

The mortality-temperature curves were generally J-shaped with a higher threshold in warmer countries (Table 1). All studies reported a positive effect of heat on mortality, which was higher in the older age groups in each of the nine studies giving estimates of populations over 65. Respiratory and cardiovascular mortality risks were higher than all-cause mortality risk, with the highest estimates seen in respiratory mortality, especially in the elderly(17,29). Heat effects were immediate or short-term (up to a few days) in all studies testing multiple lag periods(15,17,19,21,24,28). Evidence of short-term mortality displacement (harvesting effect) was found in six studies. Mortality risks were typically high for the first few days after exposure and then became negative 1-2 weeks after exposure, after which, the effect vanished(17,19,21,25,27,28). However, the harvesting effects did not account for all the excess mortality seen due to heat. The effects of geographical location on mortality displacement varied, with Baccini et al. (2008) reporting a more prolonged effect in Mediterranean than in north-continental cities, while Leone et al. (2013) detected an effect only in European cities but not in eastern-southern Mediterranean cities(17,25).

Ambient air pollution was generally found to increase heat related mortality, with NO2 reported to exacerbate the heat effect in two of the four studies in which it was modelled(22,30) and with a varying location-dependent effect in a third(17). Ozone was also found to increase mortality in two of three studies(24,28). Particulate matter tended not to have an effect on the heat-mortality relationship, with only one study claiming an increased effect(24) and three studies failing to identify any association(18,26,28).

Additional demographic and socio-economic factors found to be associated with increased heat-related mortality in the studies analysed, include: sex, the national health expenditure, unemployment rate, percent area coverage by buildings, population density, total length of roads per km^2^ within the municipality, average of mean and maximum temperature, the percentage of population born outside the European Union countries, and the percentage of early dropout from education(20,25,29,30).

### Meta-analysis

The pooled RR of mortality per unit increase in temperature above local threshold in the Mediterranean basin was found to be 1.035 (95% CI 1.028-1.041, *P*<0.0001), implying a 3.5% increase in risk of death per 1°C (Figure 2). Publication bias was assessed using a funnel plot (Supplementary Figure S4). The plot showed little evidence of bias; six studies were slightly outside the 1.96 standard error lines, but three were below the lower limits and three above the upper limits.

**Figure 2:**
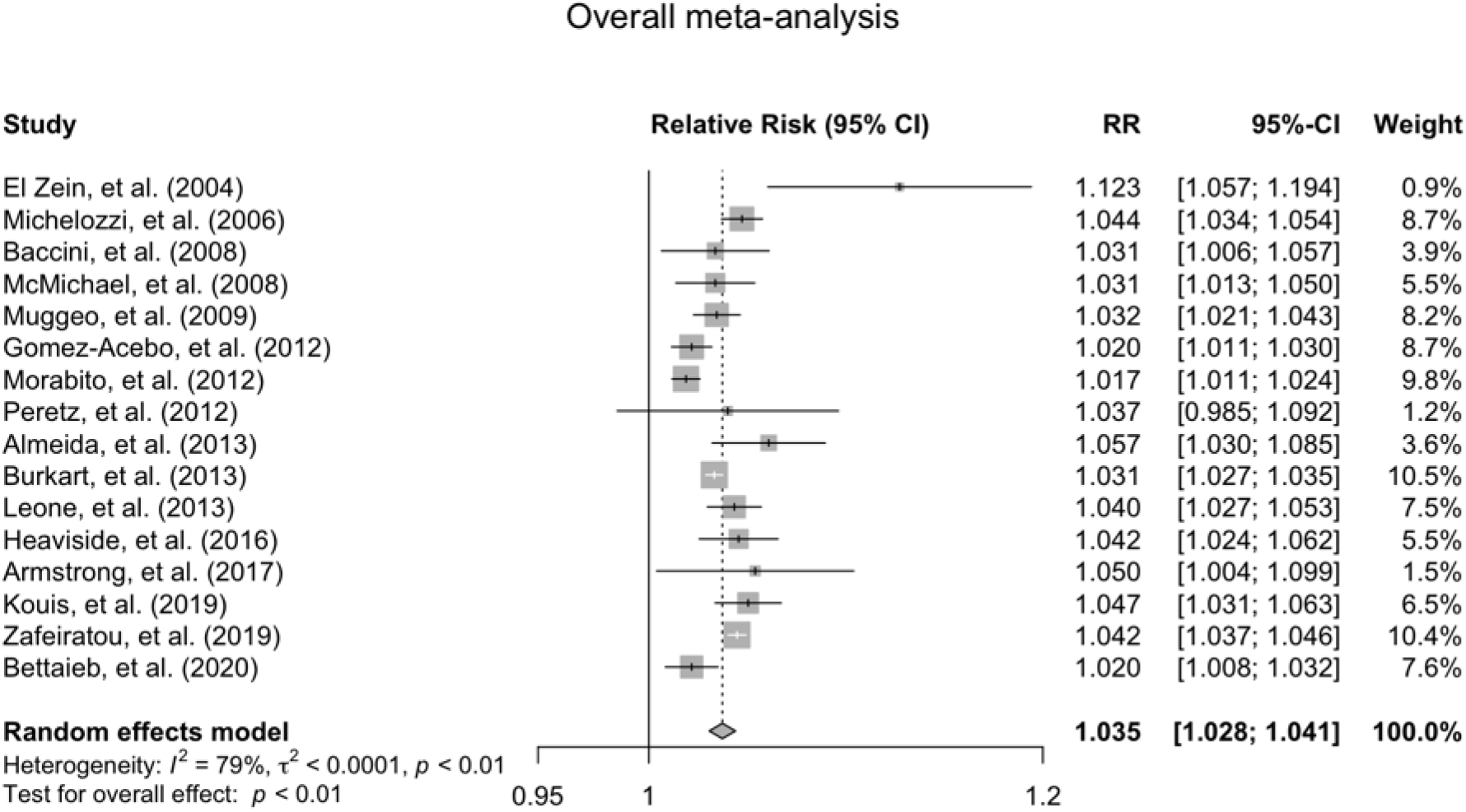
Forest plot of the meta-analysis of temperature-mortality associations for all included studies (*n*=16). Estimates are the mortality risk per 1°C increase in temperature above location-specific thresholds. RR, relative risk.

Heterogeneity was found to be moderate (I^2^=79%, Cochran’s Q=70.17). Following influence analysis (using a Baujat plot), we removed Morabito, et al. (2012) and El Zein, et al. (2004). This reduced the heterogeneity to I^2^=66.1% (Cochran’s Q=38.35), with almost no change to the overall pooled effect (RR=1.035, 95% CI 1.03-1.041, *P*<0.0001).

To account for potential factors relating to heterogeneity, we performed subgroup and meta-regression analyses.

The specific pooled RR for populations over 65 years old (nine papers; Figure 3) was similar to the all-ages RR, at 1.036 (95% CI 1.026-1.046, *P*<0.0001) per 1°C increase in temperature, with a lower heterogeneity (I^2^=57%, Cochran’s Q=18.74). Excluding El Zein, et al. (2004) decreased the RR slightly to 1.033 (95% CI 1.025-1.0404, *P*< 0.0001), and reduced the heterogeneity by around 20% (I^2^=34.3%, Cochran’s Q=10.65).

**Figure 3:**
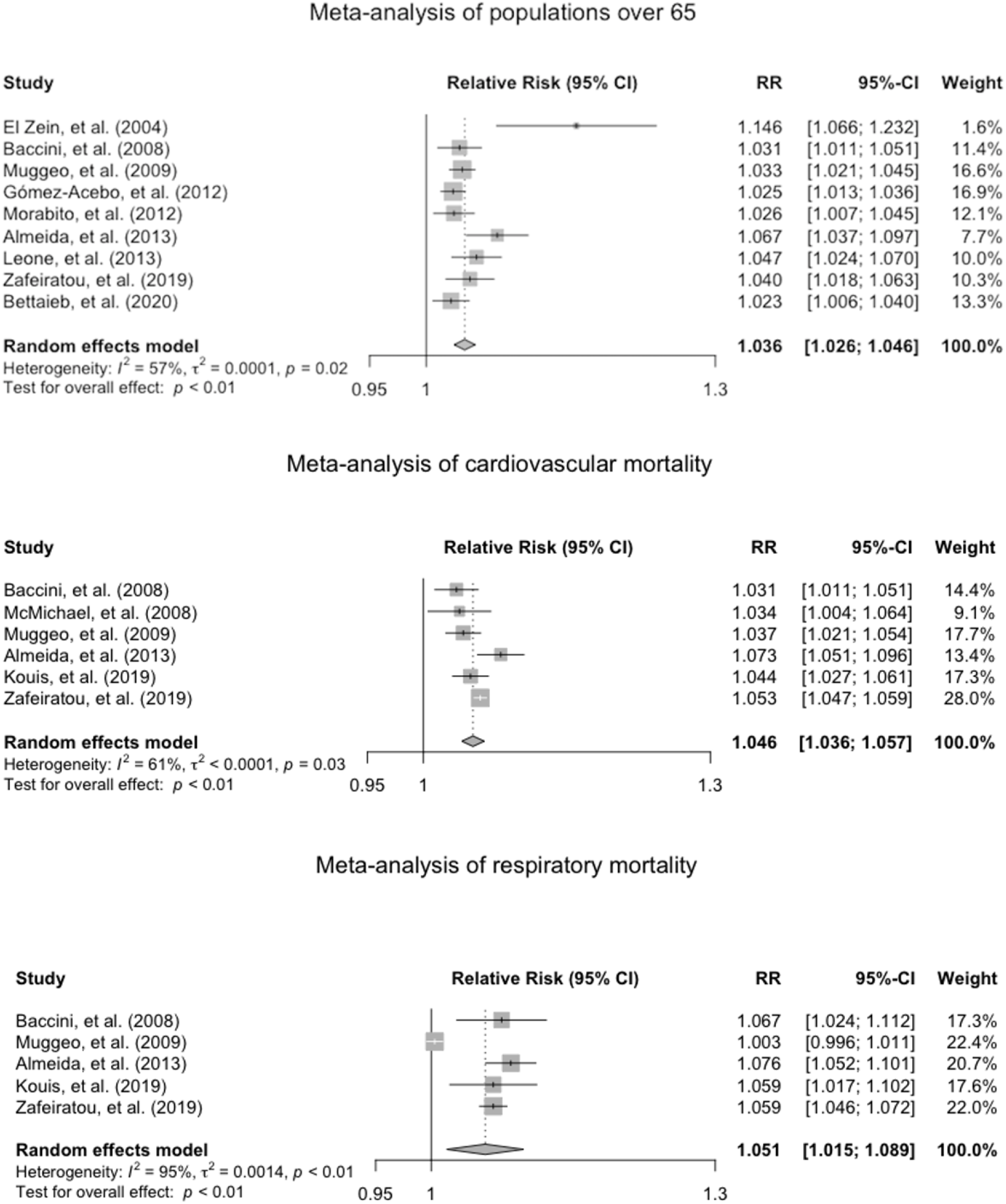
Forest plot of the meta-analysis of temperature-mortality associations in individuals over 65 years old (*n*=9), for cardiovascular mortality (*n*=6), and for respiratory mortality (*n*=5). Estimates are the mortality risk per 1°C increase in temperature above location-specific thresholds. RR, relative risk.

Meta-analysis of effect estimates for specific causes of death revealed an increased cardio-respiratory mortality risk with increased temperature (Figure 3). Cardiovascular mortality exhibited a RR of 1.046 (95% CI 1.036-1.057, *P*<0.0001) per 1°C increase in temperature above local thresholds (I^2^=60.8%, Cochran’s Q=12.76), whereas for respiratory mortality the RR was 1.051 (95% CI 1.015-1.089, *P*< 0.0058; I^2^=95.1%, Cochran’s Q=81.99). Exclusion of Muggeo, et al. (2009) dramatically reduced the heterogeneity (I^2^=0%, Cochran’s Q=1.52), and the RR of respiratory mortality rose to 1.063 (95% CI 1.052-1.074, *P*<0.0001).

### Subgroup and meta-regression analyses

The studies yielded RRs of 36 spatial units (32 city-level, 2 regional level, and 2 country-wide level), with some locations appearing more than once (in total 22 distinct spatial units across the 10 countries; Supplementary Table S2). These estimates were pooled by geographical location and by climate type.

The three major geographical locations were: western Europe (Spain, Portugal, Italy, and Slovenia), eastern Europe (Greece, Turkey, Cyprus), and the Middle East and North Africa - MENA (Lebanon, Israel, Tunisia). An association between increased temperature and mortality risk was detected in all three locations (Figure 4). There was a slight trend to higher risk of mortality in eastern Europe, with RR=1.041 (95% CI 1.034-1.048), that was somewhat lower in west Europe, RR=1.038, (95%CI 1.03-1.045), and lower still in MENA, with a RR=1.034 (95%CI 1.016-1.051), although heterogeneity remained high (I^2^=93%). Differences were detected in the pooled effect estimates of cities in various climates (*P*<0.0001), although heterogeneity remained high (I^2^=93%; Figure 5).

**Figure 4:**
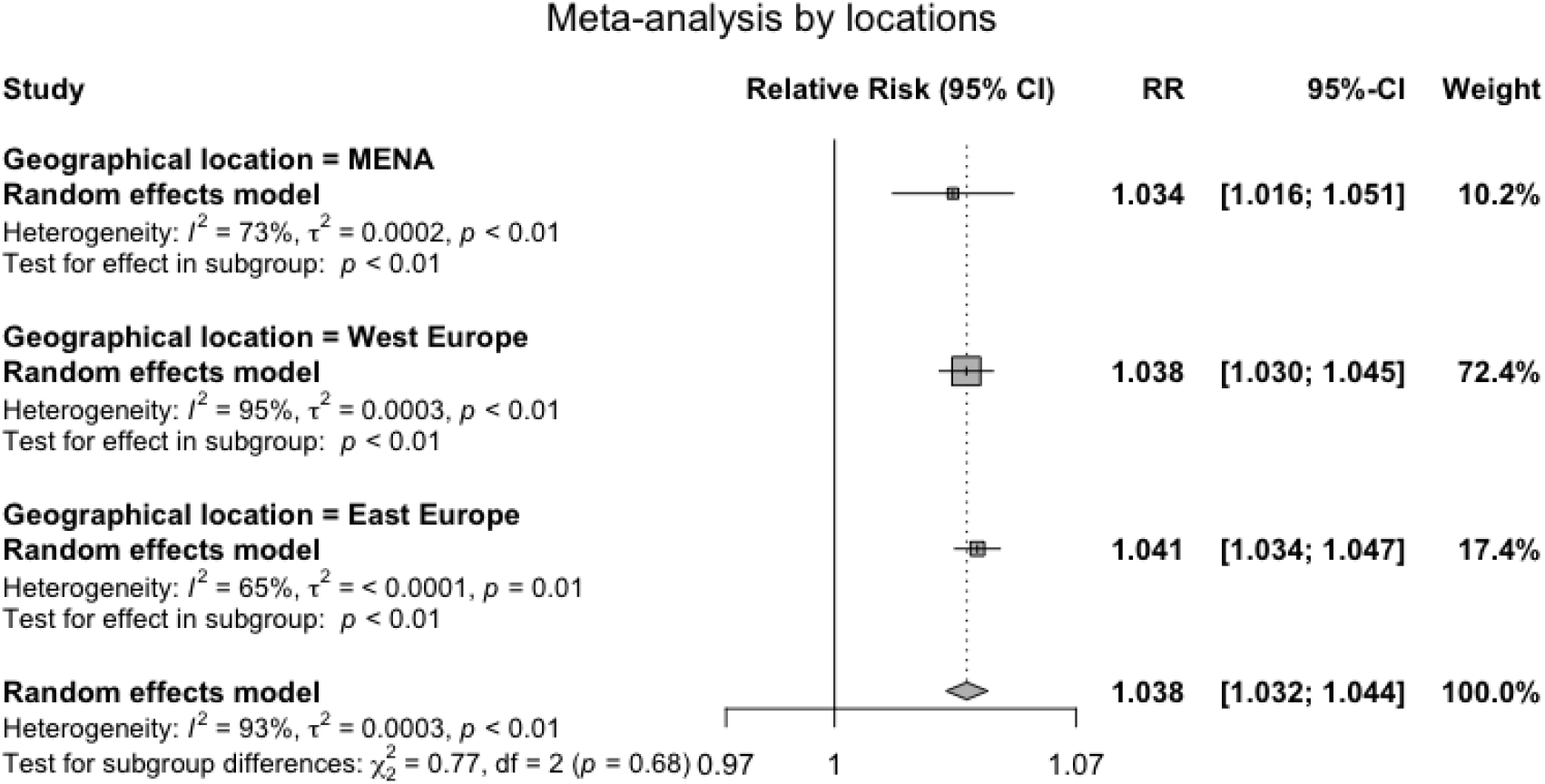
Forest plot of the meta-analysis of temperature-mortality associations stratified by geographical location: western Europe, eastern Europe (including Istanbul), and the Middle East and North Africa (MENA). Estimates are the mortality risk per 1°C increase in temperature above location-specific thresholds. RR, relative risk.

**Figure 5:**
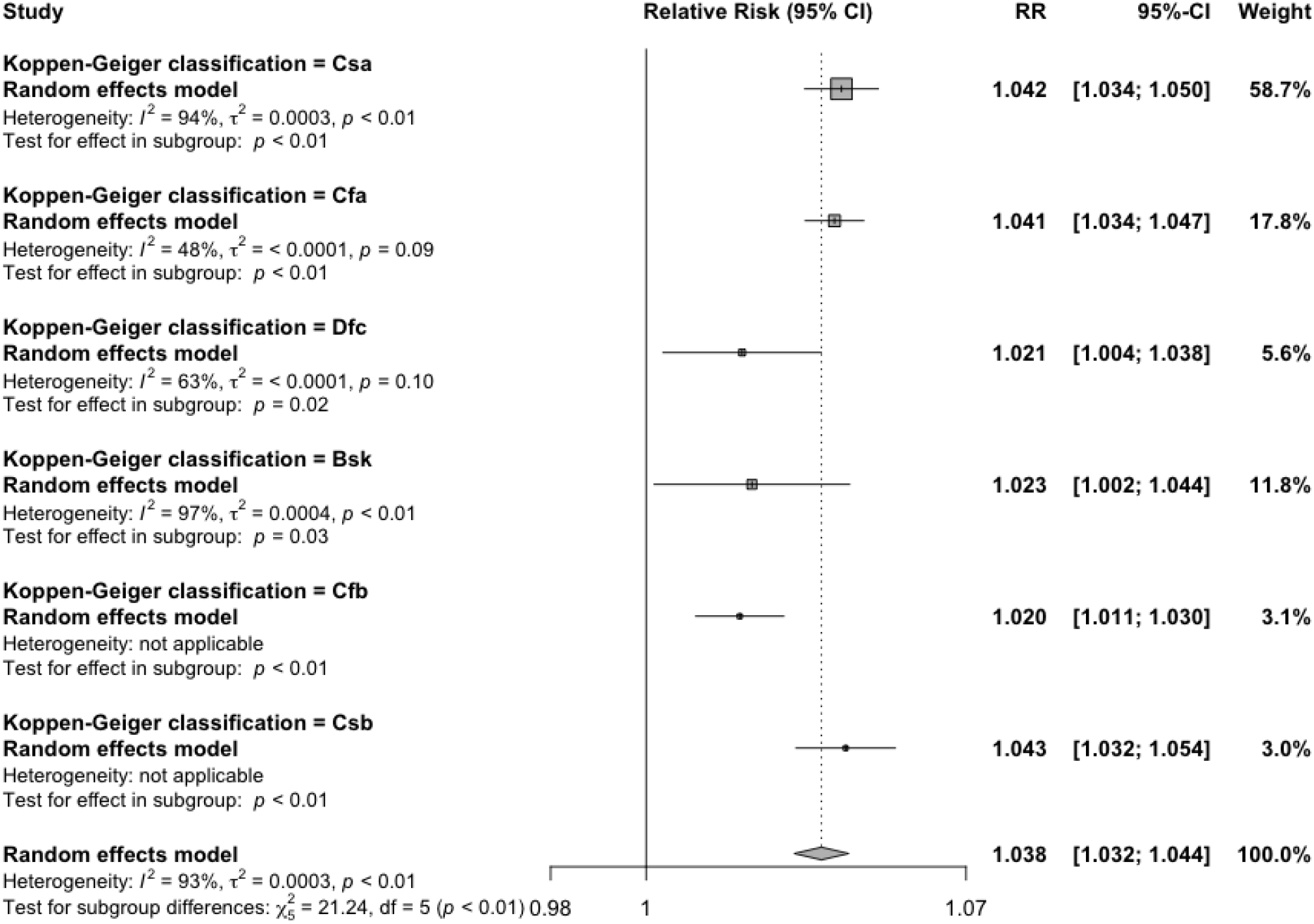
Forest plot of the meta-analysis of temperature-mortality associations stratified by Köppen-Geiger climate classification. Estimates are the mortality risk per 1°C increase in temperature above location-specific thresholds. RR, relative risk.

Meta-regression analyses by national gross domestic product (GDP) per capita, latitude, longitude, and local threshold temperatures (Supplementary Figures S5-S8) maintained the pooled RR essentially unchanged. There were slight trends towards a higher mortality risk with increasing longitude, a lower risk with increasing latitude, and a lower risk with increasing GDP per capita (albeit with heterogeneity over 90%). Similarly, there was a slight general trend for a decrease in effect estimates with increasing year of publication, but this too exhibited high heterogeneity.

Duration of study (more or less than 10 years), seasonality (all year or summer months only), and type of statistical model used in the studies did not affect the mortality risk estimates or reduce heterogeneity in additional sensitivity analyses (not shown).

## 4. Discussion

To the best of our knowledge, this is the first systematic review and meta-analysis to quantify the short-term effects of ambient heat on mortality in the Mediterranean basin. We reviewed 16 studies, all reporting statistically significant increases in mortality upon exposure to high ambient temperatures above a certain local threshold. The pooled RR for the population of all-ages in the Mediterranean basin was found to be 1.035 (95% CI 1.028-1.041, *P*<0.0001). The pooled RR for populations over 65 years old was similar, with a RR of 1.036 (95% CI 1.026-1.046, P<0001). This may be explained by the predominance of data from urban, high-income locations, where mortality is expected to involve mostly older individuals with chronic diseases.

In accordance with previous evidence, the risk of heat related mortality due to cardiovascular and respiratory causes was higher than for non-specific risks(31). In two studies, mortality from these causes was found to be particularly high in older populations(17,29). Heat thresholds in the included studies tended to be higher in warmer locations. This is in line with previous observations of long-term adaptation to heat, increasing local threshold temperatures over time, and higher thresholds in warmer locations(4,32–36).

In all studies, the short-term effects of heat were immediate, typically peaking on the day of exposure or within a few days. After such a peak, some studies showed evidence of mortality displacement (a “harvesting” effect), in which the added risk of death dropped lower than expected, returning to pre-exposure levels only after a period of 1-2 weeks. This could be due to vulnerable individuals, usually the elderly or those with cardio-respiratory diseases, whose deaths were an immediate response to the extreme conditions(37). The conclusion is that mortality displacement predominantly reflects the effect on highly susceptible individuals in the short term(38); although long-term, effects may be seen up to a year later(27). All the studies reviewed described the cumulative positive risk (total added mortality due to heat) as higher than the subsequent cumulative negative risk (displaced mortality), implying that loss of life is not all due to short-term displacement of the extremely vulnerable, but also involves subsequent substantial longer-term loss of life.

Our study has several limitations. The high between-study heterogeneity (I^2^=79%), is likely due to the different characteristics of countries in the Mediterranean basin, and varying study methodologies. Excluding two studies, as detected in an influence analysis, reduced the heterogeneity (I^2^=66.1), while maintaining a similar effect size (RR=1.035, 95%CI 1.03-1.041, *P*<0.0001). Results of subgroup analyses to account for this variation indicated that effect estimates depended on climate type (*P*<0.0001), with warmer cities tending to higher values, albeit with substantial heterogeneity (I^2^=93%). In contrast, local RR was not related to the national GDP per capita, or to the latitude and longitude of cities, local threshold temperature, or year of study publication. Regarding the inter-study differences in exposure measures, apparent temperature effects were previously reported to closely resemble those of mean daily temperature adjusted for humidity, and various temperature metrics were found likely to produce similar results(2,39). Similarly, our sensitivity analyses indicated that different exposure measures did not substantially reduce heterogeneity or alter the pooled relative risk, nor did the inclusion of seasonality, type of statistical model, or length of time-series. Hence, some differences between the studies remain unaccounted for, in part due to the lack of a gold-standard approach to estimating the effects of heat on mortality.

### Conclusion

Our results provide significant evidence for an increased risk of short-term mortality due to ambient heat exposure in the Mediterranean basin, particularly for populations >65 years, and for cardiovascular and respiratory causes of death. As climate change induces increasing temperatures and extreme weather events, it is imperative that the scientific community adopts a more unified approach to quantification of heat and the accompanying detrimental effects. Nonetheless, the evident fatal effects of heat on human populations prompt an urgent need for regional mitigation strategies targeting specific populations.

## Data Availability

All data produced in the present study are available upon reasonable request to the authors

## Author contributions

TP: Analysis, writing the original draft; UO: Conceptualization, methodology, review, and editing CP: Conceptualization, methodology, review, and editing.

## Conflict of interest

None declared.

## Supplementary data

Supplementary data are available at *IJE* online.

## Notes

### Competing Interest Statement

The authors have declared no competing interest.

### Funding Statement

This study did not receive any funding

### Author Declarations

This study involves only openly available human data, which can be obtained from the literature cited in the main text.

