## Supplementary Information for "The association between high ambient temperature and mortality in the Mediterranean basin: a systematic review and meta-analysis"

**Supplementary data**

**Table S1**: Köppen climate symbols and description (by levels)

| 1st | 2nd | 3rd | Description |
|---|---|---|---|
| A | | | **Tropical** |
| | f | | - Rainforest |
| | m | | - Monsoon |
| | w | | - Savannah |
| B | | | **Arid** |
| | W | | - Desert |
| | S | | - Steppe |
| | | h | - Hot |
| | | k | - Cold |
| C | | | **Temperate** |
| | s | | - Dry Summer |
| | w | | - Dry Winter |
| | f | | - Without Dry Season |
| | | a | - Hot Summer |
| | | b | - Warm Summer |
| | | c | - Cold Summer |
| D | | | **Cold (Continental)** |
| | s | | - Dry Summer |
| | w | | - Dry Winter |
| | f | | - Without Dry Season |
| | | a | - Hot Summer |
| | | b | - Warm Summer |
| | | c | - Cold Summer |
| E | | | **Polar** |
| | T | | - Tundra |
| | F | | - Frost |

**Adapted from**: Peel, M. C., Finlayson, B. L., and McMahon, T. A.: Updated world map of the Köppen-Geiger climate classification, Hydrol. Earth Syst. Sci., 11, 1633–1644, https://doi.org/10.5194/hess-11-1633-2007, 2007. *See reference for exact criteria*.

**Table S2**: Characteristics of minimal study spatial units for subgroup and meta-analyses

| Study | Country | City/region | Latitude | Longitude | Köppen - Geiger climate type | Geographical location |
|---|---|---|---|---|---|---|
| Heaviside, et al. (2016) | Cyprus | All Cyprus | 35.1 | 33.25 | Csa | East Europe |
| Baccini, et al. (2008) | Greece | Athens | 37.58 | 23.43 | Csa | East Europe |
| Leone, et al. (2013) | Greece | Athens | 37.58 | 23.43 | Csa | East Europe |
| Kouis et al. (2019) | Greece | Athens | 37.58 | 23.43 | Csa | East Europe |
| Zafeiratou et al. (2019) | Greece | Thessaloniki | 40.39 | 22.54 | Bsk | East Europe |
| Peretz, et al. (2012) | Israel | Tel Aviv | 32.04 | 34.47 | Csa | MENA |
| Leone, et al. (2013) | Israel | Tel Aviv | 32.04 | 34.47 | Csa | MENA |
| Leone, et al. (2013) | Italy | Bari | 41.11 | 16.87 | Cfa | West Europe |
| Michelozzi, et al. (2006) | Italy | Bologna | 44.3 | 11.21 | Cfa | West Europe |
| Armstrong, et al. (2017) | Italy | Italy | 42.2 | 12.29 | Csa | West Europe |
| Michelozzi, et al. (2006) | Italy | Milan | 45.28 | 9.11 | Cfa | West Europe |
| Baccini, et al. (2008) | Italy | Milan | 45.28 | 9.11 | Cfa | West Europe |
| Muggeo, et al. (2009) | Italy | Palermo | 38.07 | 13.22 | Csa | West Europe |
| Leone, et al. (2013) | Italy | Palermo | 38.07 | 13.22 | Csa | West Europe |
| Michelozzi, et al. (2006) | Italy | Rome | 41.54 | 12.3 | Csa | West Europe |
| Baccini, et al. (2008) | Italy | Rome | 41.54 | 12.3 | Csa | West Europe |
| Leone, et al. (2013) | Italy | Rome | 41.54 | 12.3 | Csa | West Europe |
| Michelozzi, et al. (2006) | Italy | Turin | 45.04 | 7.42 | Cfa | West Europe |
| Baccini, et al. (2008) | Italy | Turin | 45.04 | 7.42 | Cfa | West Europe |
| Morabito, et al. (2012) | Italy | Tuscany (region) | 43.77 | 11.24 | Csa | West Europe |
| El Zein, et al. (2004) | Lebanon | Beirut | 33.53 | 35.31 | Csa | MENA |
| Almeida, et al. (2013) | Portugal | Lisbon | 38.43 | 9.08 | Csa | West Europe |
| Burkart, et al. (2013) | Portugal | Lisbon | 38.43 | 9.08 | Csa | West Europe |
| Leone, et al. (2013) | Portugal | Lisbon | 38.43 | 9.08 | Csa | West Europe |
| Almeida, et al. (2013) | Portugal | Porto | 41.1 | 8.37 | Csb | West Europe |
| Baccini, et al. (2008) | Slovenia | Ljubljana | 46.03 | 14.31 | Dfc | West Europe |
| McMichael, et al. (2010) | Slovenia | Ljubljana | 46.03 | 14.31 | Dfc | West Europe |
| Baccini, et al. (2008) | Spain | Barcelona | 41.23 | 2.11 | Csa | West Europe |
| Leone, et al. (2013) | Spain | Barcelona | 41.23 | 2.11 | Csa | West Europe |
| Gomez-Acebo, et al. (2012) | Spain | Cantabria (region) | 43.18 | 3.98 | Cfb | West Europe |
| Armstrong, et al. (2017) | Spain | Spain | 40.23 | 3.43 | Bsk | West Europe |
| Baccini, et al. (2008) | Spain | Valencia | 39.28 | 0.23 | Bsk | West Europe |
| Leone, et al. (2013) | Spain | Valencia | 39.28 | 0.23 | Bsk | West Europe |
| Leone, et al. (2013) | Tunisia | Tunis | 36.48 | 10.11 | Csa | MENA |

| | | | | | | |
|---|---|---|---|---|---|---|
| Bettaieb et al. (2020) | Tunisia | Tunis | 36.48 | 10.11 | Csa | MENA |
| Leone, et al. (2013) | Turkey | Istanbul | 41.01 | 28.57 | Csa | East Europe |

**Table S3**: Study characteristics: season, statistical model, exposure, and confounders

| Author (year) | Season | Model | Lag (Days) Used | Confounders |
|---|---|---|---|---|
| El Zein et al. (2004) | All year | Poisson regression | lag 0 | **Climate**: humidity<br>**Time related variables**: day of the week, holidays, seasonality, year of death |
| Michelozzi et al. (2006) | Hot season: June to September | Poisson generalized additive model (GAM) | | |
| Baccini et al. (2008) | Hot season: April to September | Poisson generalized estimating equation model (GEE) | lag 0-3 | **Air pollution**: NO2 (lag0-1)<br>**Climate**: barometric pressure (lag0-1), wind speed<br>**Time related variables**: holidays, day of week, calendar month, long time trend |
| McMichael, et al. (2008) | All year | Poisson regression | lag 0-1 | **Air pollution**: particulate concentrations (lag 0-1)<br>**Climate**: humidity<br>**Time related variables**: day of week, public holidays, secular trends, seasonality |
| Muggeo et al. (2009) | All year | Poisson generalized additive model (GAM) | lag 0 | **Air pollution**: PM10, (lag 0-1) O3 (lag 0-1)<br>**Climate**: humidity<br>**Time related variables**: seasonality, long term trends,day of week, influenza |
| Gómez-Acebo et al. (2012) | Hot season: June to September | Poisson regression | | **Demography**: number of inhabitants<br>**Time related variables**: seasonality |
| Morabito et al. (2012) | All year | Poisson regression | lag 0-1 | **Demography**: summer population decrement<br>**Time related variables**: Year, season, day of week, holidays |
| Peretz et al. (2012) | All year | Poisson regression | lag 0-3 | **Air pollution**: NO2 (lag 0-1)<br>**Time related variables**: month, day of week |
| Almeida et al. (2013) | Hot season: April to September | Poisson generalized estimating equation model (GEE) | lag 0–3 (Tmax) | **Air pollution**: PM10, barometric pressure, wind speed<br>**Time related variables**: day of the week, holidays, calendar month, long-term time trend |

| | | | | |
|---|---|---|---|---|
| Burkart et al. (2013) | All year | Poisson generalized additive model (GAM) | lag 0-1 | **Air pollution**: PM10, O3<br>**Time related variables**: trend, year, day of the week |
| Leone et al. (2013) | Hot season: April to September | Poisson generalized estimating equation model (GEE) | lag 0-3 | **Air pollution**: NO2, barometric pressure, wind speed<br>**Climate**: mean, maximum, minimum and standard deviation of temperature and of maximum apparent temperature, mean and standard deviation of relative humidity, latitude and longitude.<br>**Socioeconomic variables**: percentage of elderly population, percentage unemployed, life expectancy at birth, infant mortality rate, population size, GDP per capita, health expenditure, hospital bed density<br>**Time related variables**: holidays, day of the week, calendar month, seasonality and long time trends |
| Heaviside et al. (2016) | Hot season: April to September | Poisson regression, DLNM | lag 0-1 | **Air pollution**: PM10<br>**Climate**: humidity<br>**Time related trends**: seasonality and long-term trends |
| Armstrong et al. (2017) | All year | Poisson regression, DLNM | lag 0-21 | **Time related trends**: seasonality, long term trend, day of week |
| Kouis et al. (2019) | All year | Poisson regression, DLNM | lag 0-1 | **Air pollutio**n: PM10, O3<br>**Climate**: humidity<br>**Time related trends**: seasonal and long-term trends |
| Zafeiratou et al. (2019) | Hot season: April to September | Poisson generalized estimating equation model (GEE) | lag 0-3 | Sociodemographic variables: population density, green space (%), density of road network, area covered by buildings, total area, immigrant population from outside EU, unemployment rate, youth unemployment rate, long term unemployment rate, population 25-64 with low secondary/upper secondary/tertiary education, early leavers from education<br>Climate: temperature from the E-OBS database<br>Time related trends: day of the week, month, seasonality and long term trends |
| Bettaieb et al. (2020) | Hot season: May to October | Poisson generalized estimating equation model (GEE) | lag 0-3 | Air pollution: NO2 (lag 0-1)<br>Time related variables: trend, calendar month, day of the week, the Ramadan period, public holidays |

In studies reporting multiple lag periods, the estimate for the shortest lag was recorded. DLNM, distributed lag nonlinear models.

* average thresholds

** pooled thresholds

**Figure S4**: Funnel plot of the log relative risk estimates and standard errors for overall meta-analysis.

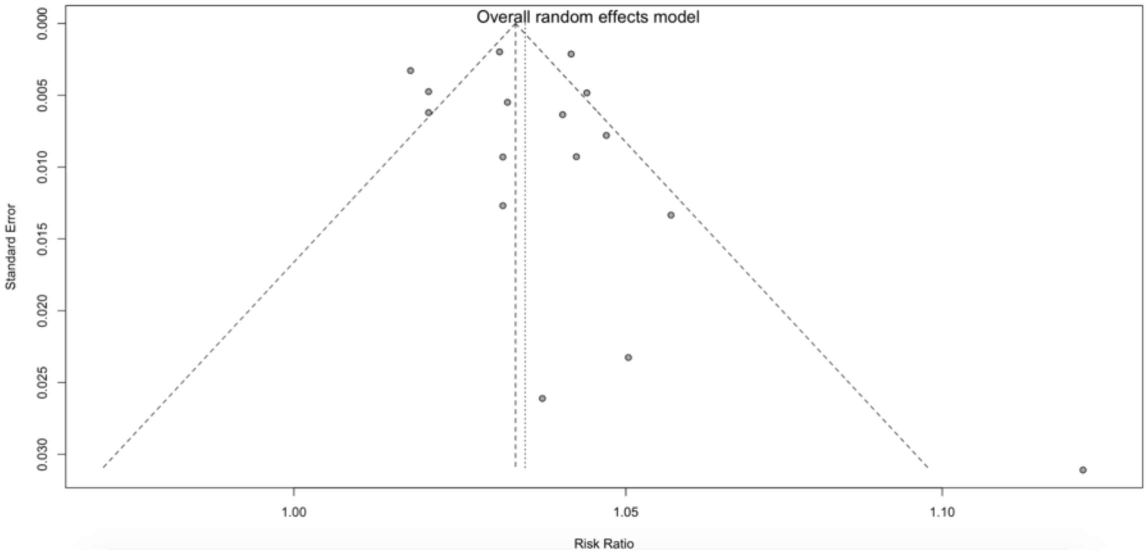


**Figure S5**: Meta-regression of temperature-mortality associations by national gross domestic product (GDP) per capita.

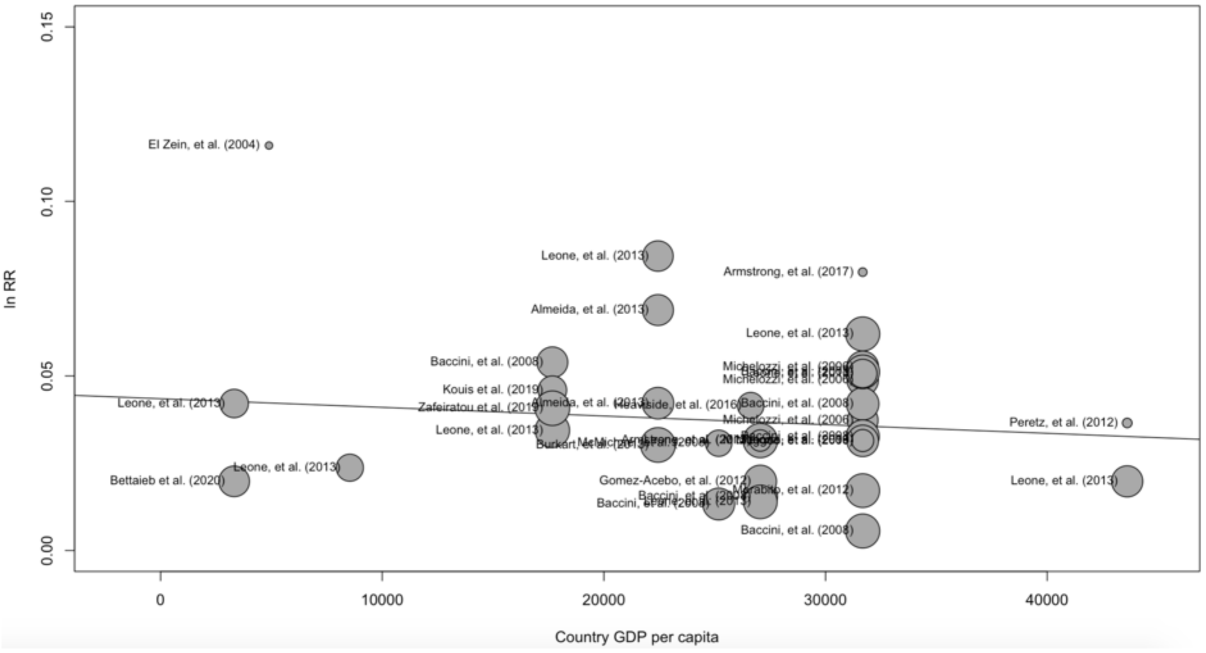

**Figure S6**: Meta-regression of temperature-mortality associations by latitude.

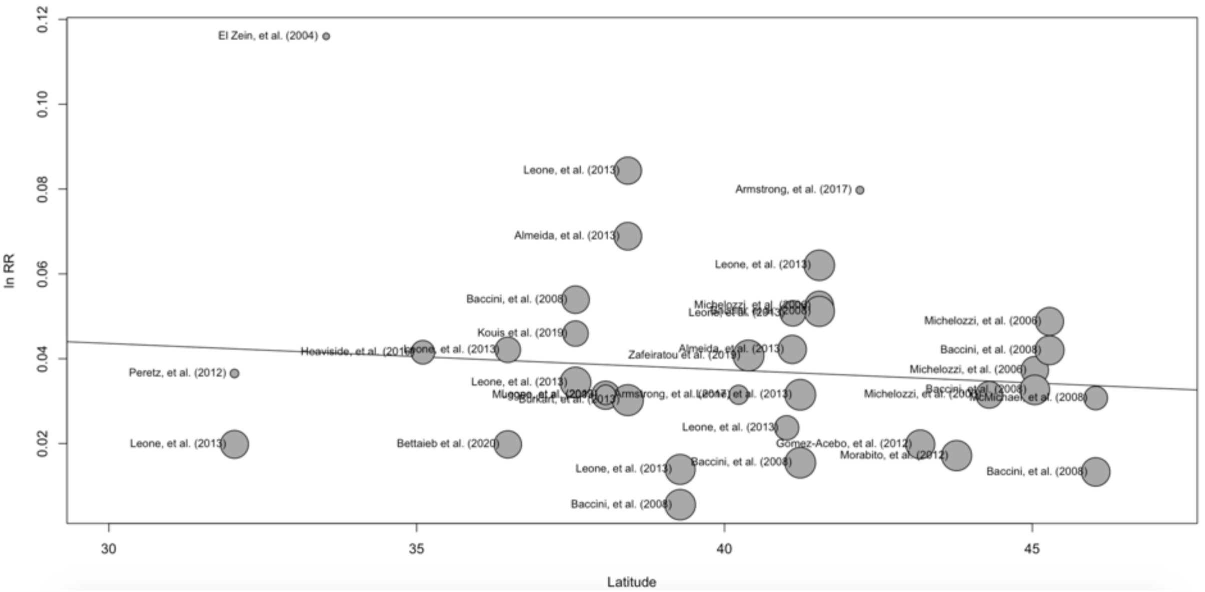


**Figure S7**: Meta-regression of temperature-mortality associations by longitude.

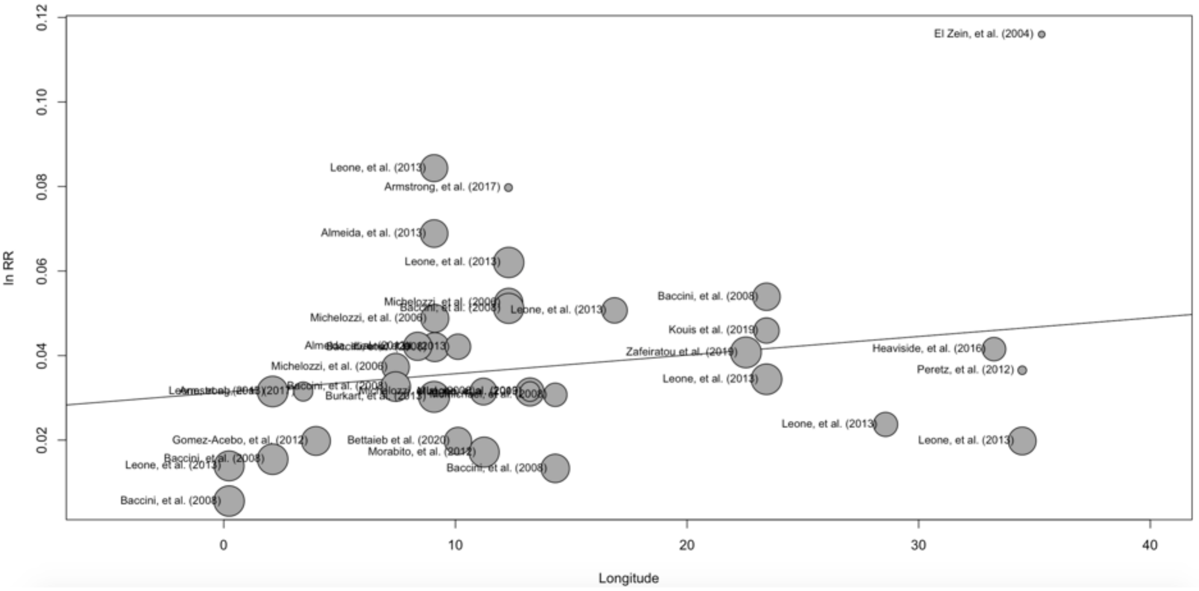

**Figure S8**: Meta-regression of temperature-mortality associations by local threshold.

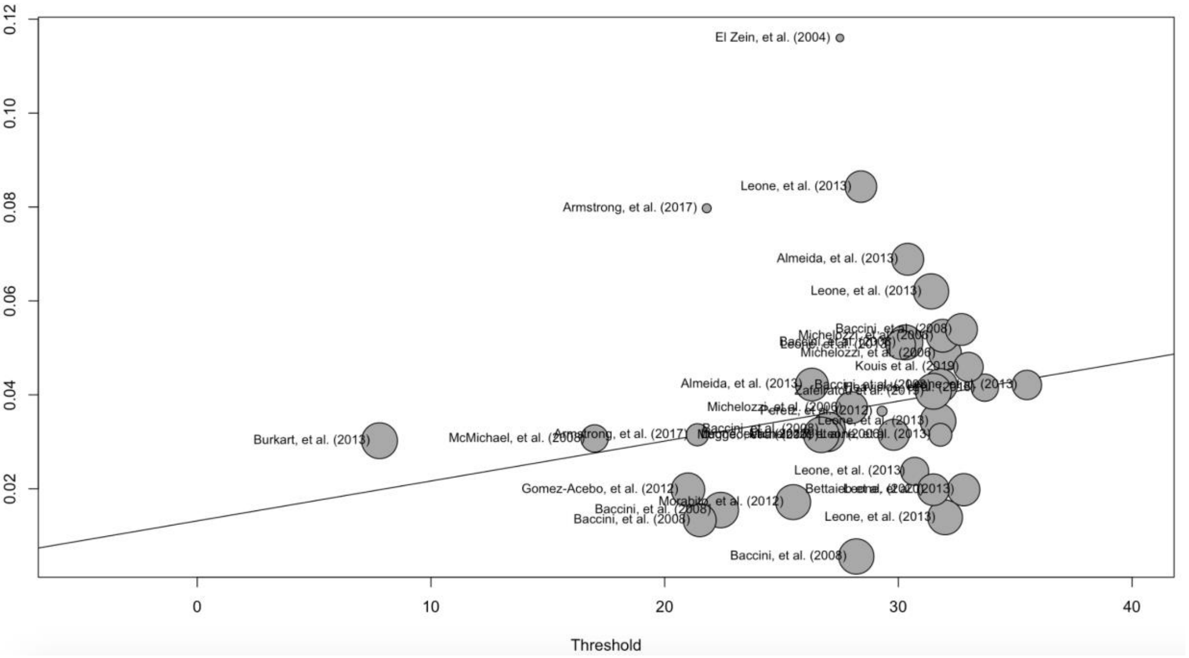
